# Effect of social distancing on COVID-19 incidence and mortality in the US

**DOI:** 10.1101/2020.06.10.20127589

**Authors:** Trang VoPham, Matthew D. Weaver, Jaime E. Hart, Mimi Ton, Emily White, Polly A. Newcomb

## Abstract

Social distancing policies were implemented in most US states as a containment strategy against severe acute respiratory syndrome coronavirus 2 (SARS-CoV-2). The effectiveness of these policy interventions on morbidity and mortality remains unknown. Our analysis examined the associations between statewide policies and objective measures of social distancing, and objective social distancing and COVID-19 incidence and mortality. We used nationwide, de-identified smartphone GPS data to estimate county-level social distancing. COVID-19 incidence and mortality data were from the Johns Hopkins Coronavirus Resource Center. Generalized linear mixed models were used to estimate incidence rate ratios (IRRs) and 95% confidence intervals (CIs) for the association between objective social distancing and COVID-19 incidence and mortality. Stay-at-home orders were associated with a 35% increase in social distancing. Higher social distancing was associated with a 29% reduction in COVID-19 incidence (adjusted IRR 0.71; 95% CI 0.57-0.87) and a 35% reduction in COVID-19 mortality (adjusted IRR 0.65; 95% CI 0.55-0.76). These findings provide evidence to inform ongoing national discussions on the effectiveness of these public health measures and the potential implications of returning to normal social activity.

## Introduction

Policies intended to increase social distancing were implemented in most US states to reduce transmission of severe acute respiratory syndrome coronavirus 2 (SARS-CoV-2). An increase in social distancing, through prohibiting social gatherings, non-essential business closures, and stay-at-home orders, may reduce disease incidence (1), but also carries personal and economic consequences. The effectiveness of the social distancing interventions implemented in the US on morbidity and mortality has not been fully described (2). This evidence is critical to inform ongoing public policy decisions and implement effective responses to future pandemics (3). We sought to examine the associations between statewide policies and objective measures of social distancing, and between objective social distancing and COVID-19 incidence and mortality in the US.

## Materials and Methods

We used nationwide, de-identified smartphone GPS data provided by Unacast to objectively estimate county-level social distancing based on: 1) change in average distance traveled (per device), 2) change in non-essential venue visitation (e.g., hair salons), and 3) the probability that two users were in close proximity (i.e., spatial distance of ≤50 m and temporal distance of ≤60 minutes) (4). Smartphone GPS devices were assigned to counties based on the longest recorded location. To calculate the change in objective social distancing for any given day, these measures were compared to the same day of the week during the pre-COVID-19 period (defined by Unacast as the four weeks prior to March 8, 2020) and scored 1-5 (higher numbers indicate increased distancing relative to the pre-COVID-19 comparator). Social distancing data were not provided for counties with a population less than 1,000; where less than 100 smartphone devices were observed for 70% of the days during the pre-COVID-19 period; or where less than 5 non-essential venues or 100 non-essential venue visits occurred during the pre-COVID-19 period. Incidence and mortality data per county were collected from the Johns Hopkins Coronavirus Resource Center (5).

For the statistical analysis, a paired t-test was used to compare objective social distancing scores before and after state stay-at-home order implementation. Generalized linear mixed models with a Poisson distribution accounting for counties nested within states were used to calculate incidence rate ratios (IRRs) and 95% confidence intervals (CIs). We used the earliest available social distancing data (February 24, 2020) as the independent variable in the incidence and mortality models. Restricted cubic regression splines were used to test for deviations from linearity. Similar results were observed using scaled Poisson models accounting for overdispersion. Multivariable models were *a priori* adjusted for variables associated with incidence rates or case ascertainment: county-level Hispanic ethnicity, non-white race, percent aged 50 years and older (6), percent males, median household income, population density, and obesity prevalence, and state-level cumulative COVID-19 testing rate. Covariate data were from the US Census Bureau 2018 American Community Survey, Robert Wood Johnson Foundation and University of Wisconsin Population Health Institute County Health Rankings and Roadmaps, and The COVID Tracking Project. The analysis encompassed the timeframe from February 24, 2020 to April 29, 2020, when some states began to reopen. All tests were two-sided and p < 0.05 was considered statistically significant. Our study did not constitute human subjects research and was considered exempt from Institutional Review Board review.

## Results

Objective social distancing data were available for 3,054 counties (94%) in all 50 states and Washington, D.C. Average social distancing prior to the first COVID-19 case, average social distancing before and after policy changes, and COVID-19 incidence rates on April 29, 2020 by county are presented in Figure 1. Forty-five states (including Washington, D.C.) implemented stay-at-home guidance. Stay-at-home orders were associated with a 35% increase in social distancing (mean score 2.01 (SD 0.74) to 2.71 (SD 0.71); p < 0.001). Each one-unit increase in social distancing was associated with a statistically significant reduction in COVID-19 incidence (adjusted IRR 0.71; 95% CI 0.57-0.87) and mortality (adjusted IRR 0.65; 95% CI 0.55-0.76) (Table 1).

**Table 1.**
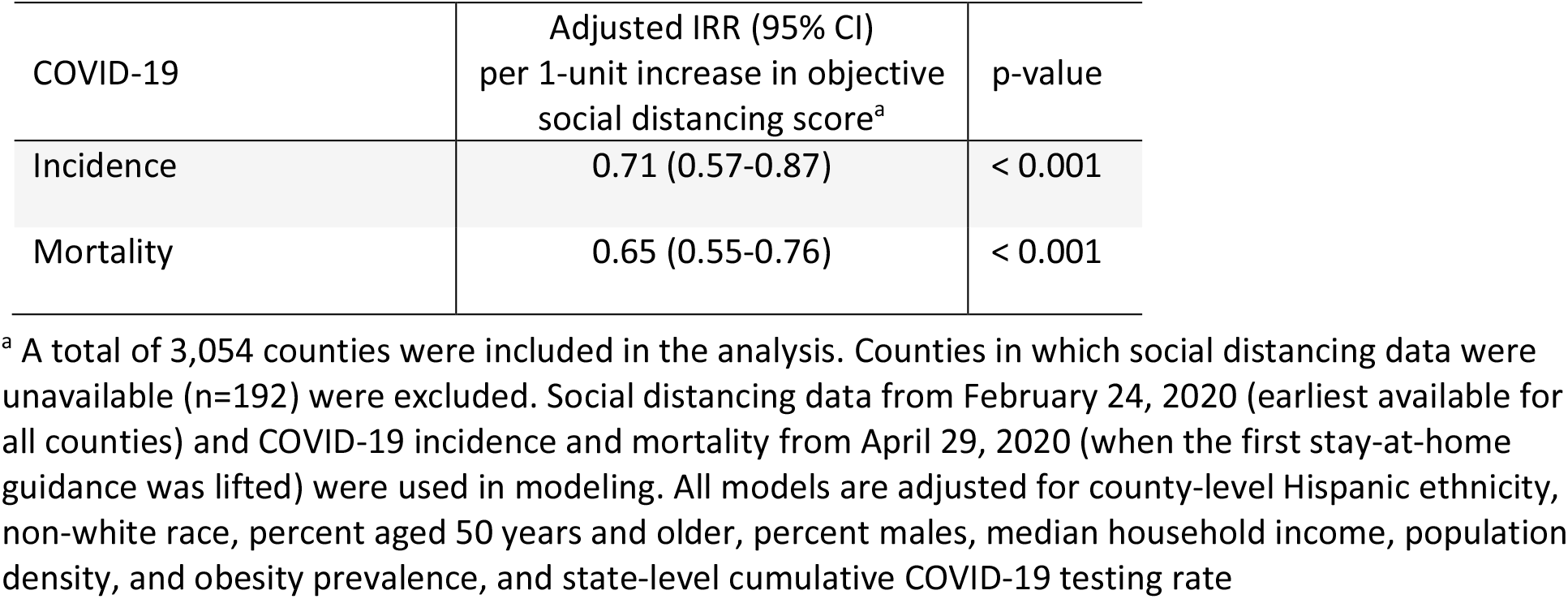
The adjusted association between county-level objective social distancing scores and COVID-19 incidence and mortality in the US

| COVID-19 | Adjusted IRR (95% CI)<br>per 1-unit increase in objective<br>social distancing score<sup>a</sup> | p-value |
| --- | --- | --- |
| Incidence | 0.71 (0.57-0.87) | $< 0.001$ |
| Mortality | 0.65 (0.55-0.76) | $< 0.001$ |

<sup>a</sup> A total of 3,054 counties were included in the analysis. Counties in which social distancing data were unavailable (n=192) were excluded. Social distancing data from February 24, 2020 (earliest available for all counties) and COVID-19 incidence and mortality from April 29, 2020 (when the first stay-at-home guidance was lifted) were used in modeling. All models are adjusted for county-level Hispanic ethnicity, non-white race, percent aged 50 years and older, percent males, median household income, population density, and obesity prevalence, and state-level cumulative COVID-19 testing rate

**Figure 1.**
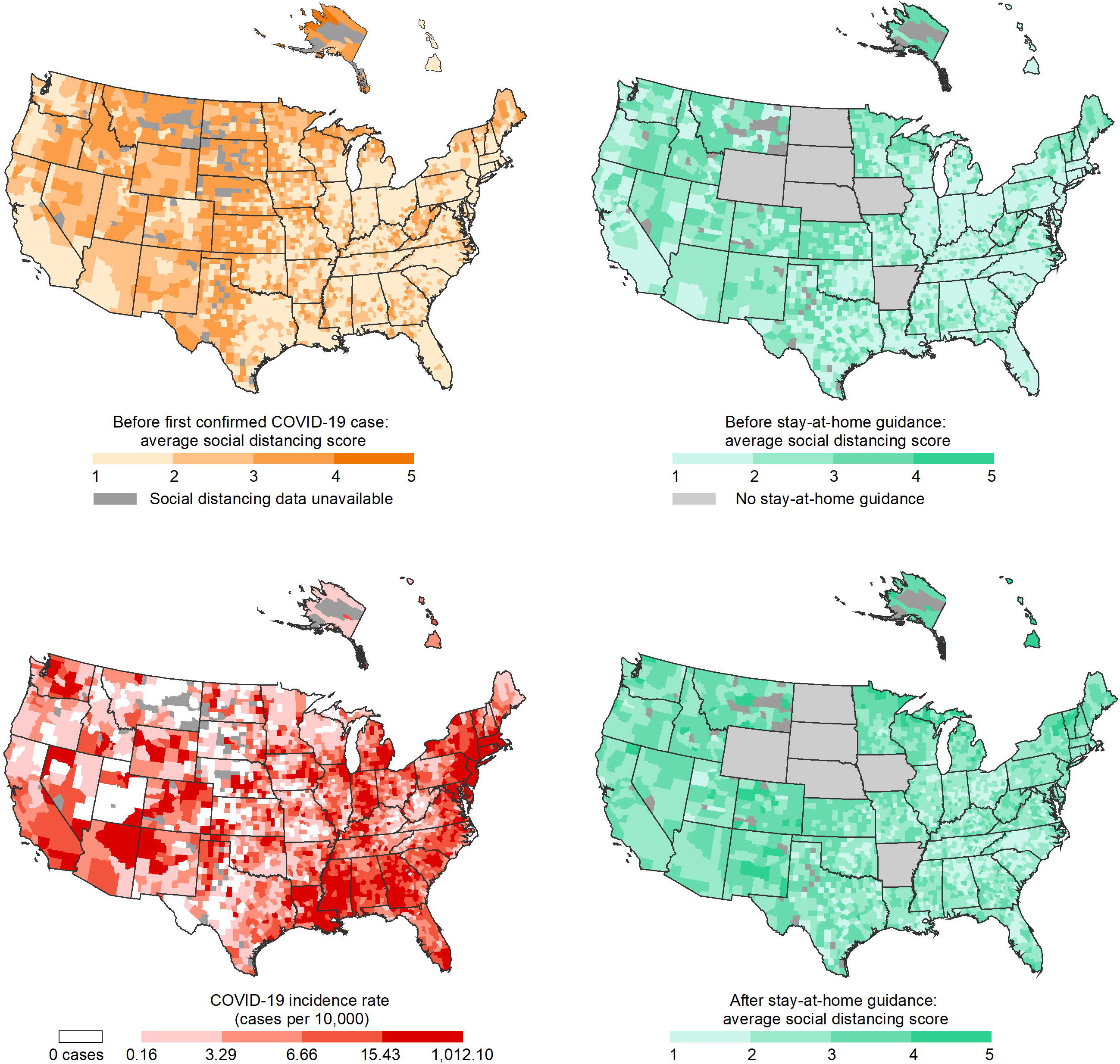
County-level objective social distancing prior to the first confirmed COVID-19 case, changes in objective social distancing before and after stay-at-home guidance, and COVID-19 incidence in the US.

Social distancing (from February 24, 2020 until the date that the first confirmed COVID-19 case occurred in each county) was mapped using the Unacast scoring scheme; average county-level social distancing was 2.03 (SD 0.85), where higher values indicate improved social distancing. Forty-five states (including Washington, D.C.) implemented stay-at-home guidance (Oklahoma and Utah enacted policies in major population centers only). Social distancing before and after stay-at-home guidance was mapped using Unacast data from February 24, 2020 to April 29, 2020. COVID-19 incidence (most current data acquired on April 29, 2020) was mapped using quartiles based on counties with >0 cases.

## Discussion

Social distancing policies implemented in states across the US resulted in meaningful behavioral change. We observed a 35% increase in objective social distancing following the implementation of state-level stay-at-home orders. This is consistent with emerging evidence using a different social distancing methodology (Google human mobility indicators) showing that state policies reduced mobility by 37% within approximately 2 weeks after their implementation (7). A separate recent effort also demonstrated that state policies were associated with a 5-10% increase in the prevalence of residents staying at home full-time (8). Our study builds on these findings by using a nationally representative dataset that objectively assessed social distancing through GPS positioning throughout the state policy implementation window, and further collected information on disease transmission and outcomes.

We found that state stay-at-home policies were successful in reducing disease. Each one-unit increase in objective social distancing was associated with a 29% reduction in COVID-19 incidence and a 35% reduction in COVID-19 mortality. Several studies have reported a downward deflection in the daily growth rate of COVID-19 cases in selected areas following these policy interventions (9, 10), though the evidence has not been entirely consistent. An analysis of bordering counties in Iowa and Illinois reported that COVID-19 incidence increased more quickly in Iowa counties, where no statewide stay-at-home order was enacted, compared to Illinois counties where a statewide stay-at-home order was put in place (11). Other studies have reported that statewide interventions have only stabilized rather than reduced disease transmission (12).

Our study was observational and we are unable to directly attribute the associations to social distancing policies. There may be residual confounding and measurement error in social distancing and outcome ascertainment, as testing was not widely accessible at the population level during our study time period.

## Conclusions

This evidence strongly suggests that policies promoting increased social distancing were beneficial. Higher social distancing was associated with marked reductions in COVID-19 incidence and mortality. These findings provide evidence to inform ongoing national discussions on the effectiveness of these public health measures and the potential implications of returning to normal social activity.

## Conflict of Interest Disclosures

The authors report no conflicts of interest.

## Funding/Support

MDW was partially supported by the National Institute for Occupational Safety and Health R01 OH011773 and the Brigham Research Institute Fund to Sustain Research Excellence. JEH was partially supported by NIH/NIEHS P30 ES000002. PAN was partially supported by NIH K05 CA152715.

## Role of the Funder/Sponsor

The funders had no role in the design and conduct of the study; collection, management, analysis, and interpretation of the data; preparation, review, or approval of the manuscript; and decision to submit the manuscript for publication.

## Disclaimer

The content is solely the responsibility of the authors and does not necessarily represent the official views of the National Institute for Occupational Safety and Health or the National Institutes of Health.

## Additional Contributions

We thank Unacast, Johns Hopkins University, US Census Bureau, Robert Wood Johnson Foundation, University of Wisconsin Population Health Institute, and The COVID Tracking Project for providing publicly available data.

## Data Availability

All social distancing, COVID-19 incidence and mortality, race/ethnicity, age, sex, socioeconomic status, population density, obesity, and COVID-19 testing files are available from public databases (https://www.unacast.com/data-for-good ; https://coronavirus.jhu.edu/ ; https://www.census.gov/programs-surveys/acs ; https://www.countyhealthrankings.org/ ; https://covidtracking.com/).

https://coronavirus.jhu.edu/

https://www.census.gov/programs-surveys/acs

https://www.countyhealthrankings.org/

https://covidtracking.com/

## Notes

### Competing Interest Statement

The authors have declared no competing interest.

### Author Declarations

Our study did not constitute human subjects research and was considered exempt from Institutional Review Board review.

## References

1 Prem K, Liu Y, Russell TW, Kucharski AJ, Eggo RM, Davies N, et al. The effect of control strategies to reduce social mixing on outcomes of the COVID-19 epidemic in Wuhan, China: a modelling study. Lancet Public Health. 2020;5(5):e261–e70.

2 Lasry A, Kidder D, Hast M, Poovey J, Sunshine G, Zviedrite N, et al. Timing of community mitigation and changes in reported COVID-19 and community mobility?four US metropolitan areas, February 26– April 1, 2020. 2020.

3 Walensky RP, Del Rio C. From Mitigation to Containment of the COVID-19 Pandemic: Putting the SARS-CoV-2 Genie Back in the Bottle. JAMA. 2020.

4 Unacast Social Distancing Dataset. 2020; https://www.unacast.com/data-for-good. Accessed version from 24 May 2020.

5 Johns Hopkins Coronavirus Resource Center. 2020; https://coronavirus.jhu.edu/. Accessed version from 24 May 2020.

6 Garg S. Hospitalization rates and characteristics of patients hospitalized with laboratory-confirmed coronavirus disease 2019—COVID-NET, 14 States, March 1–30, 2020. MMWR Morbidity and mortality weekly report. 2020;69.

7 Abouk R, Heydari B. The immediate effect of covid-19 policies on social distancing behavior in the united states. Available at SSRN. 2020.

8 Dave DM, Friedson AI, Matsuzawa K, Sabia JJ. When do shelter-in-place orders fight COVID-19 best? Policy heterogeneity across states and adoption time. National Bureau of Economic Research; 2020. Report No.: 0898-2937.

9 Courtemanche C, Garuccio J, Le A, Pinkston J, Yelowitz A. Strong Social Distancing Measures In The United States Reduced The COVID-19 Growth Rate. Health Aff (Millwood). 2020:101377hlthaff202000608.

10 Siedner MJ, Harling G, Reynolds Z, Gilbert RF, Venkataramani A, Tsai AC. Social distancing to slow the US COVID-19 epidemic: an interrupted time-series analysis. medRxiv. 2020.

11 Lyu W, Wehby GL. Comparison of Estimated Rates of Coronavirus Disease 2019 (COVID-19) in Border Counties in Iowa Without a Stay-at-Home Order and Border Counties in Illinois With a Stay-at-Home Order. JAMA Netw Open. 2020;3(5):e2011102.

12 Wagner AB, Hill EL, Ryan SE, Sun Z, Deng G, Bhadane S, et al. Social Distancing Has Merely Stabilized COVID-19 in the US. medRxiv. 2020.

